# Screening of Diabetes and Hypertension Based on Retinal Fundus Photographs Using Deep Learning

**DOI:** 10.1101/2019.12.13.19013904

**Authors:** Guangzheng Dai, Chenguang Zhang, Wei He

## Abstract

**Purpose:** The aim of this study was to use deep learning to screen for hypertension and diabetes based on retinal fundus images.

**Methods:** We collected 1160 retinal photographs which included 580 from patients with a diagnosis of hypertension or diabetes and 580 from normotensive and non-diabetic control. We divided this image dataset into (i) a development dataset to develop model and (ii) test dataset which were not present during the training process to assess model’s performance. A binary classification model was trained by fine-tuning the classifier and the last convolution layer of deep residual network. Precision, recall, the area under the ROC (AUC), and the area under the Precision-Recall curve (AUPR) were used to evaluate the performance of the learned model.

**Results:** When we used 3-channel color retinal photographs to train and test model, its prediction precision for diabetes or hypertension was 65.3%, the recall was 82.5%, the AUC was 0.745, and the AUPR was 0.742. When we used grayscale retinal photographs to train and test model, its prediction precision was 70.0%, the recall was 87.5%, the AUC was 0.803, and the AUPR was 0.779.

**Conclusions:** Our study shows that trained deep learning model based on the retinal fundus photographs alone can be used to screen for diabetes and hypertension, although its current performance was not ideal.

## INTRODUCTION

Cardiovascular diseases are the world’s biggest killers, and these diseases have remained the leading causes of death globally in the last 15 years.^1^ The key risk factors include hypertension, diabetes mellitus, dyslipidemia, cigarette smoking and obesity, which leads to progressive pathophysiological changes in various organs and tissues of the body, such as blood vessels of the brain, heart, kidney, eye. In particular, retinal vasculature, measuring 100 to 300 µm in size, attracts a lot of non-ophthalmological attention, retinal microvascular abnormalities represent an ongoing systemic microvascular damage and can be viewed directly and noninvasively, offering a unique and easily accessible “window” to study the human microcirculation in vivo. Advances in digital retinal photography and computer image analysis have now enabled more objective quantitative assessment of retinal microvascular structure and function, and may offer a potential noninvasive research tool to assess the pathophysiology of microvasculature in cardiovascular diseases and generate a cardiovascular risk prediction tool.

Extensive research on retinal microvascular phenotypes in fundus images have shown that traditional cardiovascular risk factors, such as hypertension,^2-22^ diabetes mellitus,^23-32^ dyslipidemia,^33-36^ obesity,^37-46^ drinking,^22, 33^ smoking,^33, 34, 36, 47^ can cause retinal microvascular abnormal signs to appear. These abnormal signs can be broadly divided into four groups: classic retinopathy(i.e., diabetic and hypertensive retinopathy), isolated retinopathy(e.g., retinal hemorrhage, microaneurysm, or cotton wool spot), changes in retinal vascular caliber(e.g., generalized retinal arteriolar narrowing, focal arteriolar narrowing, arteriovenous nicking), and changes in retinal vascular architecture (e.g., retinal tortuosity, fractal dimension, branching angle). These signs probably reflect systemic microvascular damage and may be an early indicator of cardiovascular diseases. There are some clinical studies supporting associations between retinal microvascular abnormalities and cerebral small-vessel disease (white matter lesions, lacunar infarcts, microbleeds),^48-53^ ischemic cerebral infarction,^54-67^ coronary atherosclerotic heart disease,^65, 66, 68-78^ congestive heart failure.^79-81^ In addition, some prospective studies suggest that retinal microvascular abnormal signs are predictive of the subsequent risk of hypertension^82-88^ or diabetes^89-93^ independently of other known risk factors.

Although a large number of studies have reflected the association between abnormal retinal microvascular signs and cardiovascular risk factors, some results were inconsistent for two reasons. Firstly, qualitative assessment of retinopathy is mainly based on the experiences of the individual and the evaluation results lacks objectivity. Secondly, there is a variety of methods of computer-assisted quantification of retinal vasculature, such as retinal vessel caliber, and thus measurements given for the same fundus image often vary. Last but not least, variations in image brightness, focus, and contrast can significantly affect the measurement of retinal vasculature. Thus this study was designed to analyze retinal image using convolutional neural networks (CNN), also known as convnets, a type of deep-learning model almost universally used in computer vision applications. One fundamental characteristic of convnets that are composed of multiple processing layers is that it can find interesting features in the training data on its own, without any need for manual feature engineering.^94^ This is especially true for problems where the input samples are very high-dimensional, like medical images (e.g., retinal fundus image, optical coherence tomography, radiographs, tissue section). It can detect subclinical and discrete features appearing below the threshold of a human observer, quantify minimal differences in feature expression and recognize patterns among large cohorts. So we trained a CNN classification model to screen for hypertension and diabetes based on retinal fundus images.

## METHODS

### Data collection

The research followed the tenets of the Declaration of Helsinki and was reviewed by the Committee on Medical Ethics of Shenyang He Eye Hospital. We collected 427 patients with a diagnosis of hypertension or diabetes and 425 normotensive and non-diabetic control subjects, who were admitted between January 2018 and December 2018 to Shenyang He Eye Hospital with eye disease. The case group include 172 males and 255 females, mean age 64.1 ± 9.6 years, whereas the control group was composed of 201 males and 224 females, mean age 53.2 ± 17.5 years.

Each patient underwent ophthalmological examination and standard assessments of cardiovascular risk factors. Retinal photographs that were centred on the macula and documented the optic disc, the macula, substantial portions of the temporal vascular arcades were taken with 45°non-mydriatic digital camera (TRC-NW300, Topcon, Tokyo, Japan) after dilation of the pupils with tropicamide phenylephrine eye drops. Hypertension was defined as systolic blood pressure greater than 140 mm Hg, diastolic blood pressure above 90 mm Hg, or use of antihypertensive medication during the previous 2 weeks. Diabetes mellitus was defined as a fasting blood glucose concentration above 7.0 mmol/L, a non-fasting value of more than 11.1 mmol/L, or a self-reported history of treatment for diabetes. Exclusion criteria were: poor dilation or ocular media opacities so that part of or the entire retinal vessel is almost indiscernible (ie, cataracts with high-severity opacity of lens, vitreous hemorrhage); any other previous or coexisting ocular disease that could affect the retinal vasculature (ie, glaucoma, central or branch retinal artery occlusion, central or branch retinal vein occlusion).

One thousand one hundred and sixty retinal photographs that include 580 from the case group and 580 from the control group. We divided this imaging datasets into a development dataset (1000 photos) to develop our models and test dataset (160 photos) which were not used during the training process to assess our model’s performance. The development dataset consist of two components: 500 images from the patients with diabetes or hypertension and 500 images form the normotensive and non-diabetic control subjects. The development dataset was divided into a training dataset (70%) and a validation dataset (30%) which was a random subset of the development dataset and was not used to train the model parameters, but was used as a small evaluation dataset for tuning the model. The test dataset consist of two components: 80 images from the patients with diabetes or hypertension and 80 images form the normotensive and non-diabetic control subjects.

### Model development

The JPEG content was decoded to RGB grids of pixels at a size of 2048×1536 and convert these into floating-point tensors. For pre-processing, auto-cutting was employed to minimize black border around the field of view and yields square images, of dimension1496×1496, in order to scale the images without distortion. All images were resized to 256×256, and rescaled pixel values (between 0 and 255) to the [0, 1] interval. To highlight the retinal vasculature, two image enhancement methods, gamma correction and contrast limited adaptive histogram equalization, were conducted to enhance the image contrast (Fig. 1). In addition, another dataset was generated by converting all 3-channel color images to the grayscale ones (Fig. 1).

**Figure 1.**
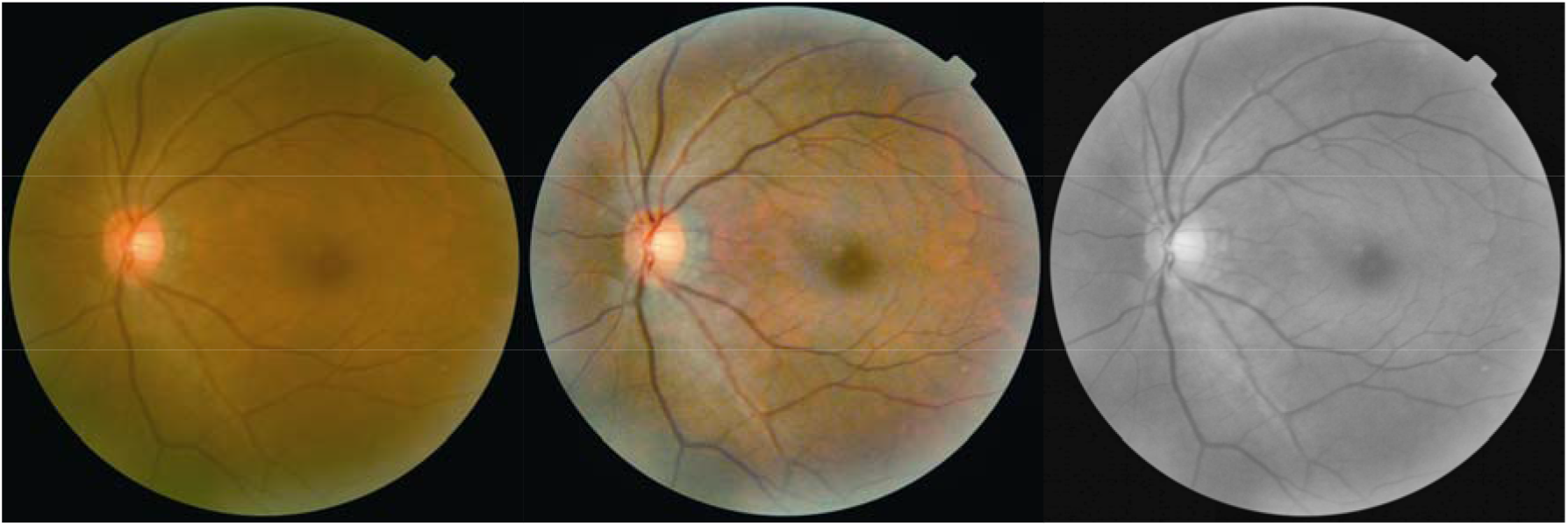
Right: original image; Middle: Enhanced image; Left: grayscale image

To avoid overfitting, data augmentation was adopted to generate more data from existing samples. The development dataset expands 1000 to 9000 through randomly extracting 9 patches of 224×224 from each 256×256 image, and each patch holds most of the vascular structure. During the training process, the training dataset via a number of random transformations, which include flipping half the images horizontally or vertically, rotating pictures within 45 degrees and translating pictures vertically or horizontally, yielding a number of believable-looking images, while the validation dataset were not augmented. Thus, at each iteration, the model never saw the exact same picture twice. This helps expose the model to more aspects of the data and generalize better.

Although data augmentation techniques were used, the inputs the model sees are still heavily inter-correlated, because they come from a small number of original images—you can’t produce new information, you can only remix existing information. As such, this may not be enough to completely get rid of overfitting. To further combat overfitting, a common and highly effective approach to deep learning on small image datasets was to use a pretrained network that is a saved network which was previously trained on a large dataset, typically on a large-scale image-classification task. The features learned by the pretrained network can prove useful for many different computer-vision problems, even though these new problems may involve completely different classes than those of the original task. In this study, deep residual network, derived from the championship model on the ImageNet Large Scale Visual Recognition Challenge of 2015(ILSVRC 2015) classification task, which are substantially deeper than deep convolutional neural networks used previously and can gain accuracy from considerably increased depth, was used for training and classification^95^. Because this study is binary classification problem, we’ll use a 2-way softmax classifier instead of the final layer with 1000 outputs and a softmax activation of the original model. We fine-tuned the classifier and the last convolution layer of resnet50, while all the other layers were be frozen, because layers higher up encode more-specialized features that need to be repurposed on new problem.

The mini-batch stochastic gradient descent (SGD) was used to optimize the trainable parameters of the resnet50. The learning rate was initialized at 0.0001 and successively decreased to 1×10^−8^ of the original per iteration, and the maximum number of iterations was 500. Momentum was used to addresses two issues with SGD : convergence speed and local minima.

### Evaluating the model

In a test phase, in order to keep the image size consistent with cropped pictures during the training process, all testing images were resized to 225×225 and randomly extracted 1 patch of 224×224. To evaluate performances of the learned model on screening for diabetes and hypertension based on fundus, we used precision, recall, the area under the ROC (AUC), the area under the Precision-Recall curve (AUPR) on separate test dataset which were not present in the development dataset.

## RESULTS

First we used pre-processed 3-channel color retinal photographs to train and test model. Kept a record of how well the model did at each epoch and plotted the loss and accuracy of the model over the training and validation data during training (figure 2). The training accuracy increased gradually with every iteration until it reached nearly 90.3%, while training loss kept decreasing until it reached nearly 0.241. The training curves were closely tracking the validation curves. The model trained at 52th epoch performed best on the test dataset with accuracy of 69.4%, and 75.7% on train dataset, 74.4% on validation dataset. On the test dataset, its prediction precision for diabetes or hypertension was 65.3%, and the recall for diabetes or hypertension was 82.5%. The model achieved an AUC of 0.745, and an AUPR of 0.742. (Table 1)

**Table 1.** The performance of the model to screen for diabetes and hypertension on different dataset

|  | Color retinal photographs | Grayscale retinal photographs |
| --- | --- | --- |
| Accuracy | 69.4% | 75.0% |
| Precision | 65.3% | 70.0% |
| Recall | 82.5% | 87.5% |
| AUC | 0.745 | 0.803 |
| AUPR | 0.742 | 0.779 |

AUC: the area under the ROC; AUPR: the area under the Precision-Recall curve

**Figure 2.**
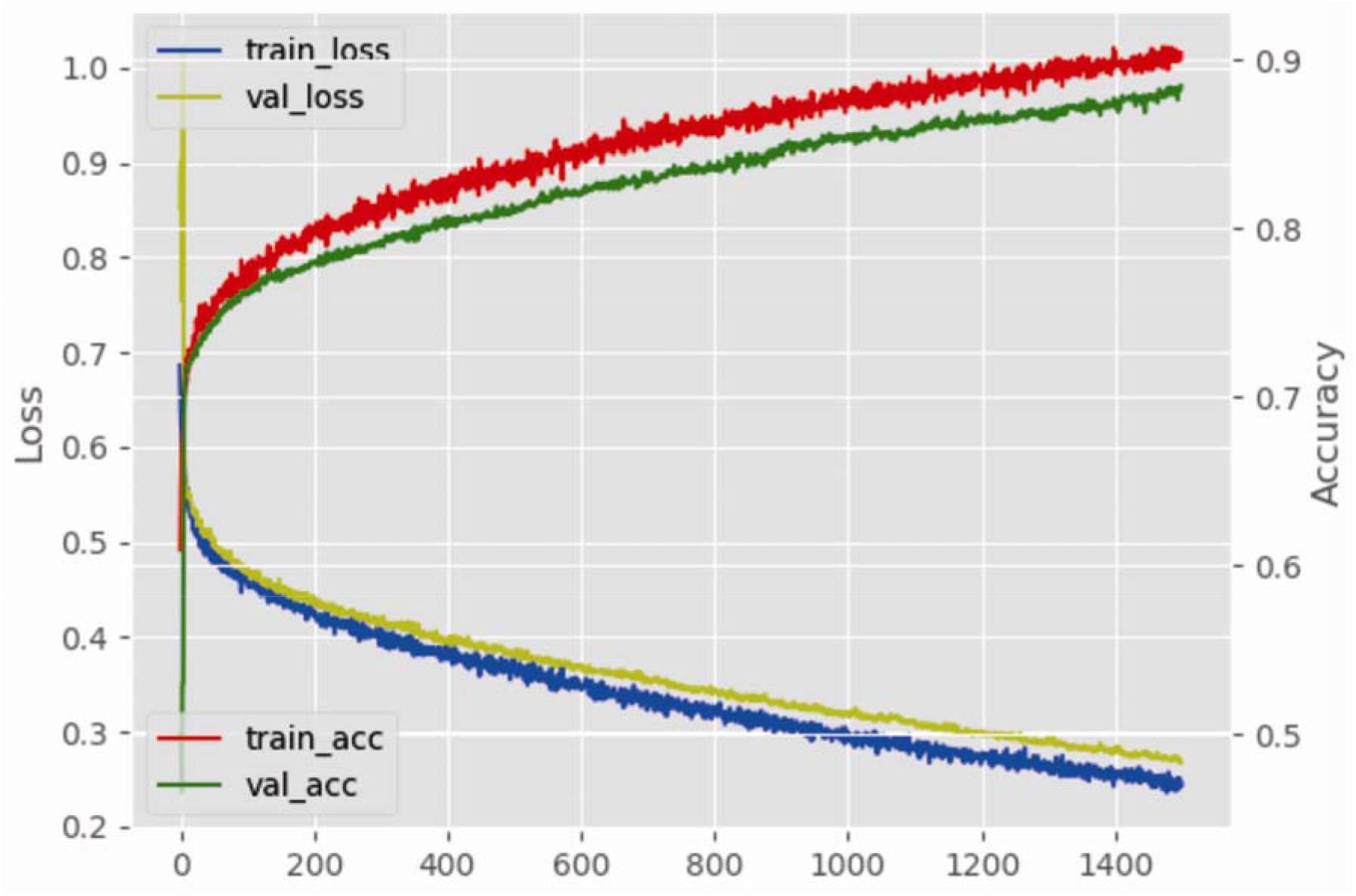
Training process on color retinal photographs

Next, we used pre-processed grayscale retinal photographs to train and test model. Kept a record of how well the model did at each epoch and plotted the loss and accuracy of the model over the training and validation data during training (figure 3).The training accuracy increased gradually with every iteration until it reached nearly 86.5%, while validation loss kept decreasing until it reached nearly 0.317. The training curves were closely tracking the validation curves. The model trained at 191th epoch performed best on the test dataset with an accuracy of 75.0%, and 77.9% on train dataset, 76.9% on validation dataset. On the test dataset its prediction precision for diabetes or hypertension was 70.0%, and the recall for diabetes or hypertension was 87.5%. The model achieved an AUC of 0.803, and an AUPR of 0.779. (Table 1)

**Figure 3.**
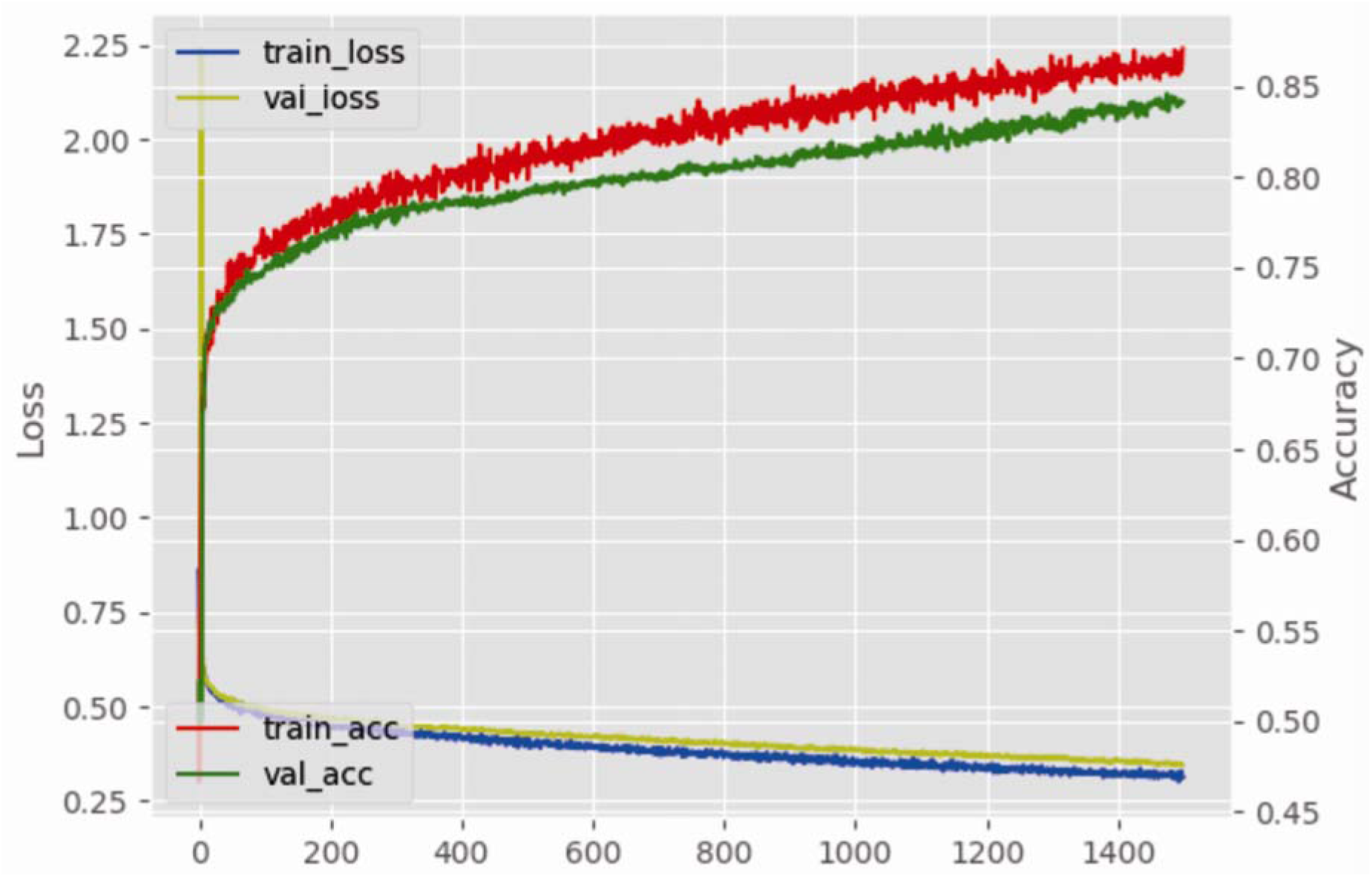
Training process on grayscale retinal photographs

There are many factors that affect the prediction accuracy of the model on separate test data samples, such as the hyper-parameters of the model including learning rate and momentum, the number of frozen layers, the number of patches randomly extracted from full image, and the size of and the proportion of patch to full image. Here are the optimal result, the others are not listed.

## Discussion

Our study show that an already trained deep learning model based on the retinal fundus photographs alone can be used to screen for diabetes and hypertension. While not ideal, these results suggests that retinal images form the patients with diabetes or hypertension has distinguishing pathological features that are likely to generalize to other new images. Substantial prior research suggests that hypertension is associated with narrower retinal arterioles,^2, 3, 5, 9-11^ decreased retinal vascular fractal dimension,^3, 17, 19-21^ less retinal arteriolar tortuosity^3, 18^ and diabetes has relation to wider retinal vessel,^23-25, 96^ decreased retinal vascular fractal dimension,^31^ more retinal arteriolar tortuosity.^30^ A deep neural network model can detect these subclinical morphological vascular changes appearing below the threshold of a human observer.

Although we use strategies for mitigating overfitting and maximizing generalization, such as data augmentation and fine-tuning, on test data, the model eventually started over-fitting after a certain number of iterations. At the beginning of training, optimization and generalization were correlated: the higher the accuracy on training data, the higher the accuracy on test data. But at 53th epoch the model trained based on color retinal photographs begin to overfit and obtain increasingly worse results on test data which it has never seen before, while based on grayscale retinal photographs it begin to overfit at 193th epoch and the gap between validation accuracy and test accuracy was narrower. So the ideal model is one that was trained at 192 epochs using grayscale retinal photographs, and at this time it stands right at the border between underfitting and overfitting. One reason for this outcome is that we had little training data available. Another possible reason could be that the fundus photograph contains a lot of additional information besides retinal microvasculature and the model learns representations that are specific to the training data but that doesn’t generalize to data outside of the training set. By converting the image to gray scale image, the additional information other than retinal vessel was compressed to the maximum extent and therefore the model trained using grayscale retinal photographs could achieve superior generalization power. To prevent a model from learning misleading or irrelevant patterns found in the training data, the best solution is to get more training data. Yet, the acquisition cost of medical image is higher and annotations must be performed by clinically qualified personals, and thousands of training images are usually beyond scope of biomedical research. Next, we construct an automated segmentation models for the retinal vessel segmentation task based on convolutional neural networks and only retained vessel morphology information. By implementing this methodology, a model can only learn retinal vascular pathological features from training dataset, thereby furthest mitigate overfitting.

Although the model’s performance on screening for diabetes and hypertension is not so ideal, our study suggest that it has certain feasibility that screening people with cardiovascular risk factors based on retinal images using deep learning. Cardiovascular risk factors, which include hypertension, diabetes mellitus, dyslipidemia, obesity and so on, lead to characteristic changes of retinal vascular morphology in fundus photograph. In the future, we will assess large databases of subclinical cardiovascular patients who often have multiple cardiovascular risk factors and use deep learning technique to train screening models based on retinal photographs. So that people at high risk of cardiovascular disease may be discovered and treated at an early stage.

## Data Availability

All data referred to in the manuscript are fully available without restriction

https://doi.org/10.5061/dryad.qrfj6q5bb

## Notes

### Competing Interest Statement

The authors have declared no competing interest.

### Funding Statement

The work received no specific funding for this work.

